# A multiplex qRT-PCR assay for detection of Influenza A and H5 subtype targeting new SNPs present in high pathogenicity avian influenza Canadian 2022 outbreak strains

**DOI:** 10.1101/2023.12.13.23298992

**Authors:** Tracy D. Lee, Frankie Tsang, Kathleen Kolehmainen, Natalie A. Prystajecky, Agatha N. Jassem, John R. Tyson

## Abstract

H5N1 is a highly pathogenic avian Influenza A subtype that has been known to also infect mammalian hosts and presents a potential public health risk. To address and mitigate the affects of new SNPs, found in recent Canadian outbreaks, on diagnostic detection we developed two qRT-PCR assays by modifying current probes to match sequences detected in the east and west coast of Canada. These assays were multiplexed with a third qRT-PCR assay targeting the M segment, allowing streamlined detection of Influenza A and subtyping for H5. This three-plex qRT-PCR was validated by assessing analytical specificity, limit-of-detection, precision, and accuracy.

## Introduction

Wild aquatic birds are the main hosts of the influenza A virus and typically include subtypes H5, H7, and H9 which are characterized by the presence of specific hemagglutinin (HA) proteins on the viral surface (1). Although transfer of avian subtypes to humans is rare, the highly pathogenic avian influenza (HPAI) subtype H5N1 has been found to cause infection in humans, with the first outbreak documented in Hong Kong in 1997 (2). There is a growing understanding for the need of a One Health approach, that is a coordinated and collaborative approach that encompasses the health of people, animals, and our shared environment (3). As such, existing assays targeting influenza A should address outbreaks and circulating strains in animals that could pose a risk to human health. Furthermore, acquisition of new mutations can compromise the ability of PCR-based typing assays to perform correctly. Rapid diagnosis using these assays is critical for individual case management and public health responses.

In 2022, HPAI infections in wild birds and commercial flocks on the east and west coast of Canada were found to contain novel single nucleotide polymorphisms (SNPs) in our existing qRT-PCR H5 subtyping targets. This led to a shift in Ct values (see **Supplementary Table S1**) compromising the assay function and prompting a need for modification of our assays. Two modified H5 qRT-PCR typing assays targeting HA (termed H5-P3 and H5-P4) were developed using 2022 sequences generated from a poultry outbreak in Newfoundland (east) and a deceased eagle found in British Columbia (west). This dual assay approach provides redundancy protection against further mutational drift with ongoing outbreaks. A third qRT-PCR assay targeting the matrix (M) gene of Influenza A (termed Flu-A M) was designed by modifying the probe sequence of a previously published assay (4). This M gene target was included to streamline detection and subtyping of suspect avian influenza virus (AIV) positive samples. The three-plex qRT-PCR assay was validated for diagnostic use by assessing analytical specificity, analytical sensitivity or limit of detection (LOD), precision, and accuracy using a panel of AIV H5 and human influenza A samples. This assay can be used to identify Influenza A and H5 subtype positivity in samples of AIV, with the potential to transition to human diagnosis if spill-over occurs.

## Material and Methods

### Assay design

Two new assays were developed to target the HA H5 segments detected in a 2022 poultry outbreak in Newfoundland (east) and from a deceased eagle found in British Columbia (west) adjusting for mutational changes identified. Primers and probes developed by the National Influenza Center at the Institut Pasteur in 2011 (5) were updated after mutational analysis of the HA sequences provided by Canadian Food Inspection Agency (CFIA) and the National Centre for Foreign Animal Disease (NCFAD) for the east and west H5 strains respectively. These sequences were provided to the BC Centre for Disease Control (BCCDC) in advance of submission to GISAID where they are now available: EPI_ISL_12968823 (east, Chicken) and EPI_ISL_15843373 (west, Bald Eagle). Both assays are able to detect the east and west sequences but target different regions of HA. For previous H5 primer and probe sequences see **Supplementary Table S2**. For comparison to modified primers and probes used in this study see **Supplementary Figs S1** and **S2**. For updated sequences used in this study, see **Table 1**. An Influenza A M segment assay developed by Fouchier et al., was modified via adjustment of the probe sequence to bring PCR cycling conditions in line with TaqMan qPCR recommendations (referred to subsequently as Flu-A M). This allowed pan subtype multiplex detection of Influenza A M segments along with H5 HA 2022 outbreak segment detection with twin redundancy using the H5-P3 and H5-P4 assays. See **Table 1** for the updated sequences and **Supplementary Figs S3** and **S4** for comparison of original and modified M segment probe.

**Table 1.**
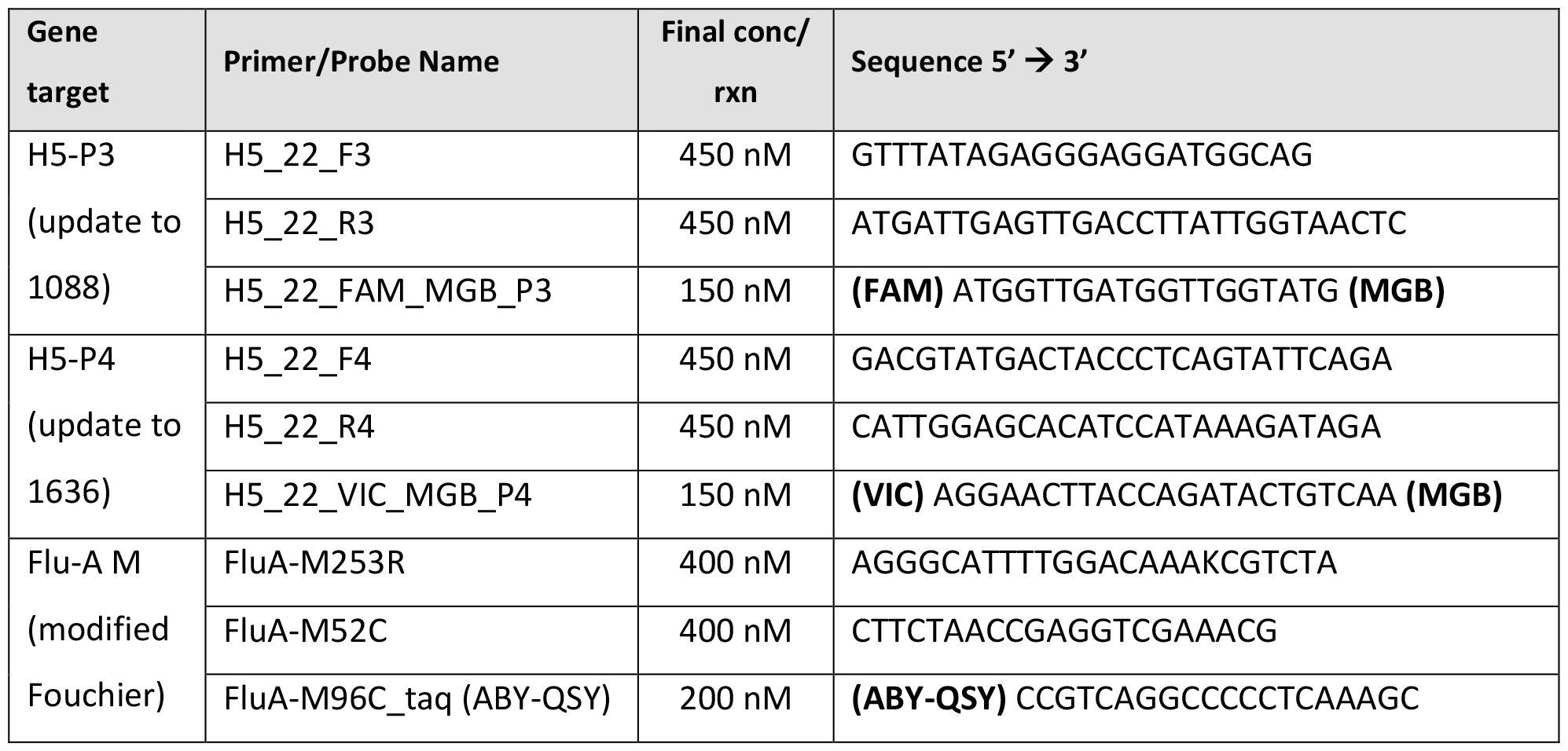
Updated Primers and Probes for Influenza A H5 and M multiplex qRT-PCR assays. Conc = concentration; rxn = reaction.

### Assay parameters

All samples were extracted using the Applied Biosystems MagMax™ Express (ThermoFisher Scientific, Canada) and 5X MagMax™-96 Viral RNA Isolation kit (Cat# AM-1836-5, ThermoFisher Scientific) per manufacturers guidelines (input volume = 200 μL, eluate volume = 60 μL). The Influenza A H5 and M multiplex qRT-PCR was performed using TaqMan™ Fast Virus 1-step Master Mix (Cat# 4444436, ThermoFisher Scientific) and Applied Biosystems QuantStudio 7 Pro (ThermoFisher Scientific). Reaction volume was 20µl and template volume was 5µl. Thresholds were set as 0.06, 0.01, and 0.05 for H5-P3, H5-P4, and Flu-A M respectively. qRT-PCR data was analyzed using Life Technologies Design & Analysis Software version 2 (ThermoFisher Scientific). Thermocycling conditions are outlined in **Table 2**.

**Table 2.** Thermocycling conditions used for multiplex qRT-PCR assay.

|  |  | Temp. (°C) | Time (Min) |
| --- | --- | --- | --- |
| Reverse Transcription |  | 50 | 5:00 |
| Initial denaturation |  | 95 | 0:20 |
| 40 Cycles | Denaturation | 95 | 0:03 |
|  | Annealing/Extension | 60 | 0:30 |

### Assay samples

Individual and pooled oropharyngeal and cloacal avian samples were collected in universal/viral transport medium (UTM/VTM) during April 2022. Human H3 and Parainfluenza samples were collected by nasopharyngeal swabs in UTM/VTM during April 2022. For specificity and accuracy experiments, a total of 30 Influenza A positive and 29 Influenza A negative samples were tested (details in **Table 3** and **Supplementary Table S3**). All samples were processed by technicians in the BCCDC Virology Lab, except for three synthetic RNA controls for H5 and human H1N1 and H3N2, which were provided by the Canadian National Microbiology Lab (NML).

**Table 3.** Summary of samples tested for analytical specificity and accuracy.

| Specimen | Count |
| --- | --- |
| Influenza A Positive | **30** |
| *AIV- H5 samples* | 15 |
| *AIV- H5 synthetic RNA* | 3 |
| *AIV- H1* | 4 |
| *Human H1* | 3 |
| *Human H3* | 5 |
| Influenza A Negative | **29** |
| *Human Parainfluenza* | 5 |
| *Influenza A Negative* | 24 |
| **Total Samples** | **59** |

Limit of detection studies (analytical sensitivity) were performed using GeneArt synthetic DNA controls (Thermofisher Scientific) and Influenza A M-Fouchier (Flu-A M) DNA gBlock controls (Integrated DNA Technologies, USA) serially diluted in IDTE with carrier RNA (sequences in **Supplementary Table S4**). A precision study was performed using 3 positive AIV samples (Samples 28, 40, and 41) at low and medium-low Ct values. Each sample was tested in triplicate over 3 separate runs.

### Confirmation of positive and negative samples

All AIV positive samples were confirmed positive for Flu A M segment using the BCCDC-developed FLUVID assay, which detects Influenza A M segment using a World Health Organization (WHO)-developed assay (5, 6). AIV-positive samples were subsequently subtyped for H5 by WGS using a previously published method (7). Non-AIV positive samples were either confirmed by Magpix, culture, or provided through NML (see **Supplementary Table S3**). Influenza A negative samples were confirmed by FLUVID. FLUVID and WGS were also used for accuracy calculations for Flu-A M and H5, respectively.

## Results

A summary of validation results is given in **Table 4**. Further description can be found in the following results sections.

**Table 4.**
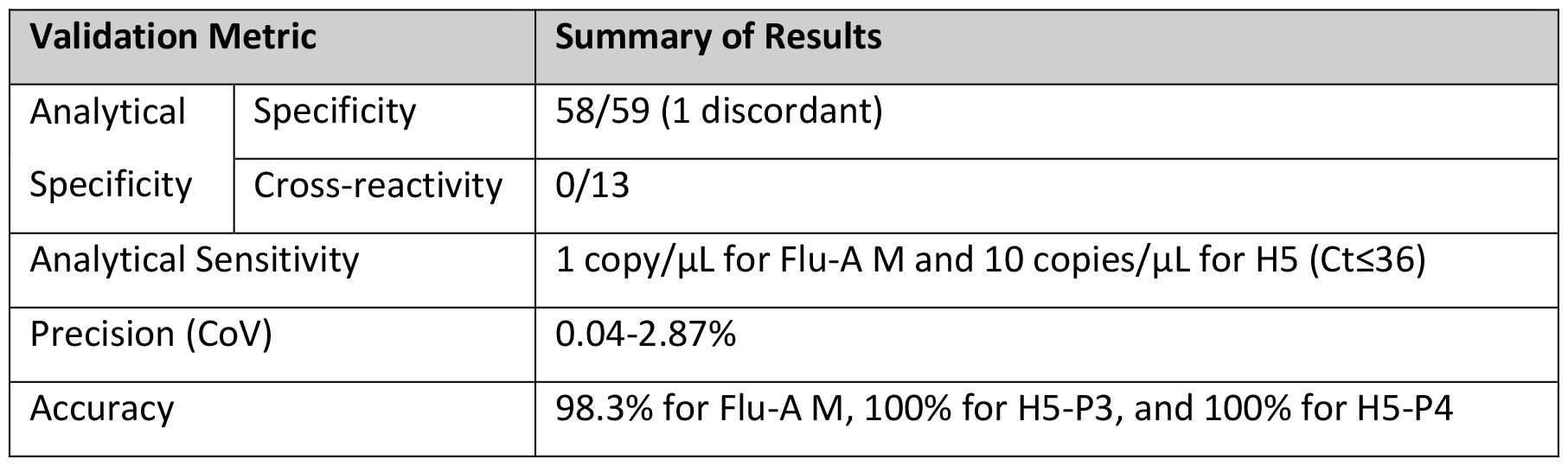
Summary of validation results. CoV = coefficient of variation.

### Analytical Specificity

Of the 59 samples tested, 1 was discordant. This sample was Flu-A M positive with a Ct >35 in this validation multiplex but was undetermined using the original Fouchier assay. Two samples were positive for Influenza (Ct >30: one AIV-H5 (matching P3 and P4 Ct) and one suspect AIV-H1 (undetermined for P3 and P4)) and were confirmed with whole genome sequencing (7). The H5 subtyping assays did not demonstrate cross-reactivity with human H1, H3, or Parainfluenza. Sample list and individual PCR results are in **Supplementary Table S3**.

### Analytical Sensitivity (LOD)

Tenfold dilutions from 10,000 to 1 copies/µL were tested in triplicate over 3 separate days to determine analytical sensitivity (for Ct values see **Supplementary Table S5**). This gave a limit of detection of 1 copy/µL or 5 copies/reaction for Flu-A M and 10 copies/µl or 50 copies/reaction for H5. Based on these results, it is recommended that an indeterminate report be issued for results that are above Ct 36.

### Precision

Ct values and precision data calculations are shown in **Supplementary Tables S6**. Coefficient of variation (CoV) for the three inter-run and intra-run experiments ranged from 0.04% - 2.87%, which is below the accepted CoV threshold of 15%.

### Accuracy

The accuracy was calculated as 98.3% for Flu-A M, 100% for H5-P3, and 100% for H5-P4 (**Supplementary Table S7**). The single sample that tested Flu-A M positive by the validation method and negative by the reference method (FLUVID) was the only discrepant sample. The Ct value for this sample was 36.1 and undetermined by the FLUVID assay (6). Of note, two additional samples were deemed concordant based on incomplete WGS data. Sample 28 (Flu-A M Ct 32.9) had a partial HA segment matching to H5 recovered and Sample 14 (Flu-A M Ct 34.8) had 1288 reads identified by Kraken2 as Influenza A along with 1154 reads mapping to segment 7 (NS1) and 921 reads mapping to segment 8 (M) (8, 9). See **Supplementary Table S3** for details.

### Interpretation criteria for qRT-PCR assay

Samples with Ct at or below 36 are interpreted as positive. If the Ct value is above 36 but below 40, then the sample can be interpreted as indeterminate. If the sample is positive or indeterminate for Flu A and positive (Ct≤36) for **both** H5-P3 and H5-P4, then the Avian H5 subtype can still be interpreted as positive. If only one of the H5 assays is positive, then the interpretation ‘unable to subtype’ should be given. If Flu A is undetermined, then the interpretation ‘Indeterminate for Flu A’ should be given regardless of the Ct value for the H5 assays.

## Discussion

Avian Influenza A (AIV) has the capacity to infect humans, leading to potentially significant impacts on human health if the highly pathogenic AIV H5 subtypes become established in the human population. Addressing circulating strains with updated, robust, and rapid assays is therefore of high importance. We developed a three-plex qRT-PCR assay targeting the M and H5 HA segments of Influenza A in response to 2022 North American AIV outbreaks. Specifically our modifications allowed detection of currently circulating North American H5 AIV strains based on sequences generated from avian samples collected in east and west Canada. This allowed for samples with a elevated Ct or undetermined using the previous H5 assays (1088 and 1636) to show robust detection with the new assays (H5-P3 and H5-P4, see **Supplementary Table S1**). One caveat is that samples were not concurrently run on both assays to assess the impact of the mutations on Ct values between them. Using relevant AIV and human Influenza A samples our updated qRT-PCR assay was evaluated for inclusivity and cross-reactivity (specificity), analytical sensitivity (LOD), precision, and accuracy. All validation parameters met internal validation criteria and, based on LOD studies, it is recommended that an indeterminate report be issued for results that are above Ct 36. As our intention is to use this assay in a One Health context, one major caveat is that endogenous controls for human and avian samples are not included to assess sample quality. As such, caution should be taken when interpreting results. Cases that are negative but of high suspicion should be confirmed by another method or laboratory. Target thresholds have been set based on data generated from the specificity tests. These thresholds should be monitored as the validation assay is deployed and further clinical or animal specimen data becomes available. Extra care should be taken to evaluate amplification and multicomponent plots of positives.

## Conclusion

A three-plex qRT-PCR assay detecting influenza A and H5 subtypes was developed to address 2022 North American HPAI outbreaks. For this particular assay, it is recommended that a positive call be made only if Ct≤36. While the novel SNPs that this assay was designed to address have only been detected in AIV samples to date, the risk for spill-over to humans makes this assay of clinical importance and further validation with human samples (if they become available) is recommended.

## Data Availability

All data produced in the present study are available upon reasonable request to the authors.

## Acknowledgements

We sincerely thank the Canadian Food Inspection Agency, National Centre for Foreign Animal Disease, Dr. Chelsea Himsworth, and Dr. Yohannes Berhane for providing sequences for use in development of this assay. We also thank all technicians and staff at the BC Wildlife AIV Surveillance Program (BC WASP) for providing samples, and the technical coordinators and technicians at the BCCDC Virology Lab for completing sample processing and extraction of all in-house samples. Furthermore we thank Kevin Kuchinski for providing the FluViewer sequencing pipeline that was used for all whole genome sequencing analyses.

## Ethics

We did not seek ethics approval as the work was a quality assurance/improvement study and did not constitute research and therefore, did not fall within the scope of REB review as per TCPS2 2022 Article 2.5 (https://ethics.gc.ca/eng/tcps2-eptc2_2022_chapter2-chapitre2.html#5).

## Notes

### Competing Interest Statement

The authors have declared no competing interest.

### Funding Statement

This study was funded through British Columbia Centre for Disease Control operational funding sources.

### Author Declarations

The Clinical Research Ethics board of the University of British Columbia waived ethical approval for this work.

## References

1. Hampson AW, Mackenzie JS. 2006. The influenza viruses. Medical Journal of Australia 185:S39–S43.

2. Chan PKS. 2002. Outbreak of Avian Influenza A (H5N1) Virus Infection in Hong Kong in 1997. Clinical Infectious Diseases 34:S58–S64.

3. 2023. One Health | CDC. https://www.cdc.gov/onehealth/index.html. Retrieved 23 October 2023.

4. Fouchier RAM, Bestebroer TM, Herfst S, Van Der Kemp L, Rimmelzwaan GF, Osterhaus ADME. 2000. Detection of Influenza A Viruses from Different Species by PCR Amplification of Conserved Sequences in the Matrix Gene. Journal of Clinical Microbiology 38:4096–4101.

5. World Health Organization. WHO information for the molecular detection of influenza viruses. https://cdn.who.int/media/docs/default-source/influenza/molecular-detention-of-influenza-viruses/protocols_influenza_virus_detection_feb_2021.pdf?sfvrsn=df7d268a_5. Retrieved 24 July 2023.

6. Eric M Hempel, Aamir Bharmal, Guiyun Li, Aileen Minhas, Ramndip Manan, Kathy Doull, Lynsey Hamilton, Branco Cheung, Michael Chan, Kingsley Gunadasa, Ron Chow, Tracy Lee, Frankie Tsang, Karen Mooder, Natalie Prystajecky, Agatha Jassam, Linda M N Hoang. Accepted. Prospective, Clinical Comparison of Self-Collected Throat-Bilateral Nares Swabs, Saline Gargle and Health Care Provider Collected Nasopharyngeal Swabs among Symptomatic Outpatients with Potential SARS-CoV-2 Infection. JAMMI. Accepted.

7. Mitchell PK, Cronk BD, Voorhees IEH, Rothenheber D, Anderson RR, Chan TH, Wasik BR, Dubovi EJ, Parrish CR, Goodman LB. 2021. Method comparison of targeted influenza A virus typing and whole-genome sequencing from respiratory specimens of companion animals. J Vet Diagn Invest 33:191–201.

8. Kevin Kuchinski. 2022. FluViewer (Version 0.1.0, test branch, git commit id:4f3be8a848cd698a519d89569029d63c041825d). github: https://github.com/BCCDC-PHL/FluViewer and https://github.com/BCCDC-PHL/FluViewer_nf with database: https://github.com/BCCDC-PHL/FluViewer/blob/main/FluViewer_db.fa.gz.

