## Supplemental Tables S1-7 for "A multiplex qRT-PCR assay for detection of Influenza A and H5 subtype targeting new SNPs present in high pathogenicity avian influenza Canadian 2022 outbreak strains"

### Supplementary Tables

**Table S1.** qRT-PCR runs comparing previous H5 assay (H5 1088 and H5 1636) to redesigned H5 assay (H5-P3 and H5-P4). Targets not tested in these runs (by assay) are shaded in dark gray. Und = Undetermined.

| Sample Name | H5 1088 | H5 1636 | H5-P3 | H5-P4 |
|---|---|---|---|---|
| 2022-04-14 Run | | | | |
| Sample 27 | 31.41 | Und | 21.17 | 20.15 |
| FLU A H1N1 control | Und | Und | Und | Und |
| Extraction negative control | Und | Und | Und | Und |
| Gblock PCR control for H5 1088 and H5 1636 (~200 copies/uL) | 29.01 | 28.42 | | |
| No template control (NTC) | Und | Und | Und | Und |
| Gblock PCR control for H5 EAST (~2000 copies/uL) | | | 26.98 | 25.25 |
| Gblock PCR control for H5 WEST (~2000 copies/uL) | | | 27.39 | 25.62 |
| 2022-04-21 Run | | | | |
| Sample 23 | 27.23 | Und | 21.18 | 19.94 |
| Sample 4 | 38.30 | Und | 31.33 | 30.43 |
| Sample 13 | Und | Und | Und | Und |
| Sample 20 | Und | Und | Und | Und |
| Sample 14 | Und | Und | Und | Und |
| Sample 26 | Und | Und | Und | Und |
| FLU A H1N1 control | Und | Und | Und | Und |
| Extraction negative control | Und | Und | Und | Und |
| Gblock PCR control for H5 1088 and H5 1636 (~200 copies/uL) | 29.79 | 28.36 | | |
| NTC | Und | Und | Und | Und |
| Gblock PCR control for H5 EAST (~2000 copies/uL) | | | 27.22 | 25.07 |
| Gblock PCR control H5 WEST (~2000 copies/uL) | | | 27.66 | 25.62 |

**Table S2.** Primers and Probes used in previous H5 qPCR assays

| Gene Target | Primer/Probe Name | Final Concentration /reaction (nM) | Sequence 5' → 3' |
|---|---|---|---|
| H5 1088** | H5-1063Fwd | 800 | TTTATAGAGGGAGGATGG |
| | H501162Rv | 800 | GAGTGGATTCTTTGTCTG |
| | H5-1088 (MAX/ZEN) | 200 | TGGTAGATGGTTGGTATGGG |
| H5 1636** | H5-1544Fw | 800 | CCGCAGTATTCAGAAGAAGC |
| | H5-1683Rv | 800 | AGACCAGCYAYCATGATTGC |
| | H5d-1638(+) FAM/ZEN | 200 | AGTGCTAGRGAACTCGCMACTGTAG |
| ** National Influenza Center (Norther-France), Institut Pasteur, Paris[1] | | | |

[1] World Health Organization. WHO Information for molecular diagnosis of influenza virus in humans-update (2011 Aug)

**Table S3.** Samples tested for Specificity & Accuracy. Und = Und. Samples not tested by detection methods or WGS are listed as NA.

| Sample | **Detection** | | | | **Validation** | | | **WGS HA** | **Previous ID** | **Previous ID method** | **Comment** |
|---|---|---|---|---|---|---|---|---|---|---|---|
| | **BCCDC FLUVID** | **Flu-A M** | **H5-P3** | **H5-P4** | **Flu-A M (0.05)** | **H5-P3 (0.06)** | **H5-P4 (0.1)** | | | | |
| SAMPLE 1 | Und | NA | NA | NA | Und | Und | Und | NA | - | - | |
| SAMPLE 2 | Und | NA | NA | NA | Und | Und | Und | NA | - | - | |
| SAMPLE 3 | Und | NA | NA | NA | Und | Und | Und | NA | - | - | |
| SAMPLE 4 | 27.36 | 28.08 | 31.33 | 28.88 | 28.73 | 27.81 | 28.92 | H5 | - | - | |
| SAMPLE 5 | Und | NA | NA | NA | Und | Und | Und | NA | - | - | |
| SAMPLE 6 | Und | NA | NA | NA | Und | Und | Und | NA | - | - | |
| SAMPLE 7 | Und | NA | NA | NA | **36.12** | Und | Und | NA | - | - | below LOD |
| SAMPLE 8 | Und | NA | NA | NA | Und | Und | Und | NA | - | - | |
| SAMPLE 9 | Und | NA | NA | NA | Und | Und | Und | NA | - | - | |
| SAMPLE 10 | Und | NA | NA | NA | Und | Und | Und | NA | - | - | |
| SAMPLE 11 | Und | NA | NA | NA | Und | Und | Und | NA | - | - | |
| SAMPLE 12 | Und | NA | NA | NA | Und | Und | Und | NA | - | - | |
| SAMPLE 13 | 32.72 | 30.89 | Und | Und | 31.48 | Und | Und | **H1** | - | - | H1N1 |
| SAMPLE 14 | 34.75 | 34.10 | Und | Und | 34.79 | Und | Und | **Failed** | - | - | Failed WGS, M and NS1 segment identified via de novo assembly in geneious 9.1.8 and contigs blasted |
| SAMPLE 15 | Und | NA | NA | NA | Und | Und | Und | NA | - | - | |
| SAMPLE 16 | Und | NA | NA | NA | Und | Und | Und | NA | - | - | |
| SAMPLE 17 | Und | NA | NA | NA | Und | Und | Und | NA | - | - | |
| SAMPLE 18 | Und | NA | NA | NA | Und | Und | Und | NA | - | - | |
| SAMPLE 19 | Und | NA | NA | NA | Und | Und | Und | NA | - | - | |
| SAMPLE 20 | 34.21 | 33.41 | Und | Und | 34.07 | Und | Und | **H1** | - | - | replicate Flu A M Ct 34.86; H1N1 |
| SAMPLE 21 | Und | NA | NA | NA | Und | Und | Und | NA | - | - | |
| SAMPLE 22 | Und | NA | NA | NA | Und | Und | Und | NA | - | - | |
| SAMPLE 23 | 17.05 | 17.61 | 21.18 | 18.39 | 18.07 | 17.63 | 18.43 | H5 | - | - | |
| SAMPLE 24 | Und | NA | NA | NA | Und | Und | Und | NA | - | - | |
| SAMPLE 25 | Und | NA | NA | NA | Und | Und | Und | NA | - | - | |
| SAMPLE 26 | 30.10 | 27.92 | Und | Und | 28.56 | Und | Und | **H1** | - | - | H1N1 |
| SAMPLE 27 | 17.54 | 17.84 | 21.17 | 20.15 | 18.29 | 18.12 | 18.60 | H5 | - | - | |
| SAMPLE 28 | NA | 32.00 | 32.73 | 32.73 | 32.86 | 30.00 | 32.80 | H5 | - | - | partial H5 segment recovered from WGS data |
| SAMPLE 29 | NA | 20.36 | 23.92 | 20.51 | 21.42 | 20.83 | 21.05 | H5 | - | - | |
| SAMPLE 30 | NA | 23.48 | 27.05 | 23.35 | 24.21 | 24.24 | 24.09 | H5 | - | - | |
| SAMPLE 31 | Und | NA | NA | NA | Und | Und | Und | NA | - | - | |
| SAMPLE 32 | Und | NA | NA | NA | Und | Und | Und | NA | - | - | |
| SAMPLE 33 | Und | NA | NA | NA | Und | Und | Und | NA | - | - | |

| | | | | | | | | | | |
|---|---|---|---|---|---|---|---|---|---|---|
| SAMPLE 34 | Und | NA | NA | NA | Und | Und | Und | NA | - | - |
| SAMPLE 35 | NA | 26.10 | 33.50 | 25.96 | 26.67 | 27.62 | 26.62 | H5 | - | - |
| SAMPLE 36 | NA | 16.54 | 19.46 | 16.48 | 17.36 | 16.56 | 16.95 | H5 | - | - |
| SAMPLE 37 | NA | 22.13 | 24.45 | 21.94 | 22.73 | 21.19 | 22.18 | H5 | - | - |
| SAMPLE 38 | NA | 17.14 | 21.29 | 17.86 | 18.00 | 18.21 | 18.35 | H5 | - | - |
| SAMPLE 39 | NA | 19.15 | 23.23 | 19.91 | 19.70 | 20.01 | 20.34 | H5 | - | - |
| SAMPLE 40 | NA | 24.83 | 27.99 | 24.88 | 25.11 | 24.45 | 25.44 | H5 | - | - |
| SAMPLE 41 | NA | 28.86 | 32.12 | 29.22 | 28.95 | 28.75 | 29.28 | H5 | - | - |
| SAMPLE 42 | NA | 21.73 | 25.04 | 22.32 | 22.30 | 22.08 | 22.83 | H5 | - | - |
| SAMPLE 43 | NA | 26.70 | 29.51 | 26.85 | 27.10 | 26.29 | 27.29 | H5 | - | - |
| SAMPLE 44 | - | - | NA | NA | 17.40 | Und | Und | NA | H3 | Magpix |
| SAMPLE 45 | - | - | NA | NA | 31.35 | Und | Und | NA | H3 | Magpix |
| SAMPLE 46 | - | - | NA | NA | 20.57 | Und | Und | NA | H3 | Magpix |
| SAMPLE 47 | - | - | NA | NA | 24.61 | Und | Und | NA | H3 | Magpix |
| SAMPLE 48 | - | - | NA | NA | 21.30 | Und | Und | NA | H3 | Magpix |
| SAMPLE 49 | - | - | NA | NA | Und | Und | Und | NA | Parainfluenza 2 | Magpix |
| SAMPLE 50 | - | - | NA | NA | Und | Und | Und | NA | Parainfluenza 3 | Magpix |
| SAMPLE 51 | - | - | NA | NA | Und | Und | Und | NA | Parainfluenza 4 | Magpix |
| SAMPLE 52 | - | - | NA | NA | Und | Und | Und | NA | Parainfluenza 4 | Magpix |
| SAMPLE 53 | - | - | NA | NA | Und | Und | Und | NA | Parainfluenza 4 | Magpix |
| Flu A Calif | - | - | NA | NA | 13.41 | Und | Und | NA | Flu A H1N1 | Culture |
| Flu A HK | - | - | NA | NA | 10.68 | Und | Und | NA | Flu A H3N2 | NML control |
| Flu A Hawaii | - | - | NA | NA | 8.37 | Und | Und | NA | Flu A H1N1 | NML control |
| RNA control 2.3.4.4b | - | - | - | - | 21.94 | 19.43 | 19.63 | | | |
| A/CK/BC/FAV22 8/2022 | - | - | - | - | 12.70 | 12.51 | 13.52 | | | |
| A/BAEA/BC/ OTH33-36/2022 | - | - | - | - | 10.87 | 11.72 | 11.62 | | | |

**Table S4.** H5 GeneArt String and Influenza A gBlock Sequences. Primers and probes are shaded grey. P4 is underlined and *P3 is in italics*

| Name | Sequence |
|---|---|
| GeneArt String EAST: 571bp | ATAGCAGTGACGTATGACTACCCTCAGTATTCAGAAGAAGCAAGATTAAAAAGAGAAGAAATAAGCGGAGTGAAATTAGAATCAGTAGGAACTTACCAGA<br>TACTGTCAATTTATTCAACAGCGGCAAGTTCCCTAGCACTGGCAATCATGATGGCTGGTCTATCTTTATGGATGTGCTCCAATGACGT*GTTTATAGAGGGAG*<br>*GATGGCAGGGAATGGTTGATGGTTGGTATGGGTACCATCATAGCAATGAGCAGGGGAGTGGGTACGCTGCGGACAAAGAATCCACCCAAAAGGCAATAG*<br>*ATGGAGTTACCAATAAGGTCAACTCAATCAT*ACGTGACTCAACAATTATGAAAAGTGGAGTGGAATATGGCCACTGCAACACCAAATGTCAAACCCCAGTA<br>GGAGCGATAAATTCTAGTATGCACGTTTAAACCAGAGGTTGGCACCAAAAATAGCTACTAGATCCCAAGTAAACGGGCAACGTGGAAGAATGGACTTCTTC<br>TGGACAATCTTAAAACCAGATGATGCAATCCATTTCGAGAGTAATGGAAATTTCATTGCTTGCCAGTA |
| Flu-A M gBlock: 800bp | GTTTGAGTCTGTTGCTTGGTCAGCAAGTGCTTGCCATGATGGCACCAGTTGGTTGACAATTGGAATTTCTGGCCCAGACAATGGGGCTGTGGCTGTATTGAA<br>ATACAATGGCATAATAACAGACACTATCAAGAGTTGGAGGAATAACATACTGAGAACTCAAGAGTCTGAATGTGCATGTGTAAATGGCATCCGCAGTATTC<br>AGAAGAAGCAAGACTAAAAAGAGAGGAAATAAGTGGAGTAAAATTGGAATCGATAGGAATTTACCAAATACTGTCAATTTATTCTACAGTGGCGAGTTCCC<br>TAGCACTGGCAATCATGGTAGCTGGTCTATTTTATAGAGGGAGGATGGCAGGGAATGGTAGATGGTTGGTATGGGTACCACCATAGCAATGAGCAGGGGA<br>GTGGGTACGCTGCAGACAAAGAATCCACTCGTCTTCTAACCGAGGTCGAAACGTACGTTCTTTCTATCATCCCGTCAGGCCCCCTCAAAGCCGAGATCGCAC<br>AGAGACTGGAAAGTGTCTTTGCAGGAAAGAACACAGATCTTGAGGCTCTCATGGAATGGCTAAAGACAAGACCAATCTTGTCACCTCTGACTAAGGGAATT<br>TTAGGATTTGTGTTCACGCTCACCGTGCCCAGTGAGCGAGGACTGCAGCGTAGACGCTTTATCCAAAATGCCCTATCGGCTTATAGCAAAAACCAACCAACA<br>ATTTGAGTTGATAGACAATGAATTCAATGAGGTAGAGAAGCAAATCGGTAATGTGATAAATTGGACCAGAGATTCTATAACAGAAGTGTGG |

**Table S5.** Limit of detection/analytical sensitivity Ct values. Threshold is in brackets.

| **Assay (threshold)** | **Flu-A M (0.05)** | **H5-P3 (0.06)** | **H5-P4 (0.1)** |
|---|---|---|---|
| **Sample** | **Flu-A M gBlock 1 copy/µl** | **GeneArt String EAST 10 copies/µl** | |
| **Standard Curve** | 35.18 | 37.01 | 36.22 |
| | 34.49 | 37.77 | 37.26 |
| | 35.73 | 37.32 | 36.57 |
| | 35.03 | - | - |
| | 35.85 | - | - |
| | 35.68 | - | - |
| *Intra-run average* | *35.33* | *37.37* | *36.68* |
| *SD* | *0.52* | *0.38* | *0.53* |
| *CoV* | *1.48%* | *1.01%* | *1.44%* |
| **Sensitivity 1** | 34.59 | 35.58 | 35.30 |
| | 34.79 | 35.44 | 35.64 |
| | 36.59 | 35.20 | 35.05 |
| *Intra-run average* | *35.32* | *35.41* | *35.33* |
| *SD* | *1.11* | *0.20* | *0.30* |
| *CoV* | *3.13%* | *0.55%* | *0.84%* |
| **Sensitivity 2** | 34.78 | 35.57 | 36.00 |
| | 35.79 | 36.55 | 36.78 |
| | 34.86 | 36.17 | 36.50 |
| *Intra-run average* | *35.15* | *36.10* | *36.43* |
| *SD* | *0.56* | *0.50* | *0.40* |
| *CoV* | *1.59%* | *1.37%* | *1.09%* |
| **Inter run average** | **35.28** | **36.29** | **36.15** |
| **SD** | **0.64** | **0.92** | **0.72** |
| **CoV** | **1.82%** | **2.54%** | **1.99%** |

**Table S6.** Precision tables for Flu-A M, H5-P3, and H5-P4. Threshold is in brackets.

| | **Flu-A M (0.05)** | | | **H5-P3 (0.06)** | | | **H5-P4 (0.1)** | | |
|---|---|---|---|---|---|---|---|---|---|
| | SAMPLE 28 | SAMPLE 40 | SAMPLE 41 | SAMPLE 28 | SAMPLE 40 | SAMPLE 41 | SAMPLE 28 | SAMPLE 40 | SAMPLE 41 |
| **Precision 1** | 32.36 | 27.08 | 28.60 | 29.81 | 26.61 | 28.30 | 32.45 | 27.10 | 28.94 |
| | 32.40 | 27.03 | 28.43 | 30.04 | 26.57 | 28.40 | 32.44 | 27.11 | 28.92 |
| | 32.52 | 26.82 | 28.28 | 29.79 | 26.56 | 28.31 | 32.40 | 27.04 | 28.85 |
| *Intra-run Average* | *32.43* | *26.98* | *28.44* | *29.88* | *26.58* | *28.34* | *32.43* | *27.08* | *28.90* |
| *Intra-run SD* | *0.08* | *0.14* | *0.16* | *0.14* | *0.02* | *0.05* | *0.03* | *0.04* | *0.05* |
| *CoV (%)* | *0.26%* | *0.51%* | *0.55%* | *0.47%* | *0.09%* | *0.19%* | *0.08%* | *0.14%* | *0.16%* |
| **Precision 2** | 32.52 | 27.04 | 28.41 | 29.81 | 26.61 | 28.30 | 32.45 | 27.10 | 28.94 |
| | 32.28 | 27.07 | 28.37 | 30.04 | 26.57 | 28.40 | 32.44 | 27.11 | 28.92 |
| | 32.31 | 26.90 | 28.30 | 29.79 | 26.56 | 28.31 | 32.40 | 27.04 | 28.85 |
| *Intra-run Average* | *32.37* | *27.01* | *28.36* | *29.88* | *26.58* | *28.34* | *32.43* | *27.08* | *28.90* |
| *Intra-run SD* | *0.13* | *0.09* | *0.06* | *0.14* | *0.02* | *0.05* | *0.03* | *0.04* | *0.05* |
| *CoV (%)* | *0.41%* | *0.33%* | *0.19%* | *0.47%* | *0.09%* | *0.19%* | *0.08%* | *0.14%* | *0.16%* |
| **Precision 3** | 32.99 | 27.22 | 28.50 | 29.86 | 26.91 | 28.17 | 32.39 | 27.32 | 29.47 |
| | 34.01 | 27.57 | 28.55 | 30.19 | 26.92 | 28.60 | 32.95 | 27.29 | 29.13 |
| | 34.93 | 27.88 | 28.40 | 30.24 | 26.93 | 28.41 | 33.25 | 27.24 | 28.94 |
| *Intra-run Average* | *33.98* | *27.56* | *28.49* | *30.10* | *26.92* | *28.39* | *32.86* | *27.28* | *29.18* |
| *Intra-run SD* | *0.97* | *0.33* | *0.08* | *0.21* | *0.01* | *0.22* | *0.44* | *0.04* | *0.27* |
| *CoV (%)* | *2.87%* | *1.20%* | *0.26%* | *0.69%* | *0.04%* | *0.77%* | *1.34%* | *0.14%* | *0.93%* |
| **Inter-run Average** | **32.92** | **27.18** | **28.43** | **29.95** | **26.69** | **28.36** | **32.58** | **27.15** | **29.00** |
| **Inter-run SD** | **0.93** | **0.34** | **0.11** | **0.18** | **0.17** | **0.12** | **0.31** | **0.11** | **0.20** |
| **Inter run CoV (%)** | **2.83%** | **1.24%** | **0.37%** | **0.60%** | **0.64%** | **0.42%** | **0.95%** | **0.39%** | **0.68%** |

**Table S7.** Accuracy calculations for validation assay targets

| **Flu-A M** | | *Reference Method (BCCDC FLUVID)* | |
|---|---|---|---|
| | | **Positive** | **Negative** |
| **H5 3plex** | **Positive** | 30 | 1 |
| | **Negative** | 0 | 28 |
| **H5-P3** | | *Reference Method (BCCDC Whole Genome Sequencing)* | |
| | | **Positive** | **Negative** |
| **H5 3plex** | **Positive** | 18 | 0 |
| | **Negative** | 0 | 41 |
| **H5-P4** | | *Reference Method (BCCDC Whole Genome Sequencing)* | |
| | | **Positive** | **Negative** |
| **H5 3plex** | **Positive** | 18 | 0 |
| | **Negative** | 0 | 41 |
