## Supplemental Figures S1-4 for "A multiplex qRT-PCR assay for detection of Influenza A and H5 subtype targeting new SNPs present in high pathogenicity avian influenza Canadian 2022 outbreak strains"

**Supplementary Figures**

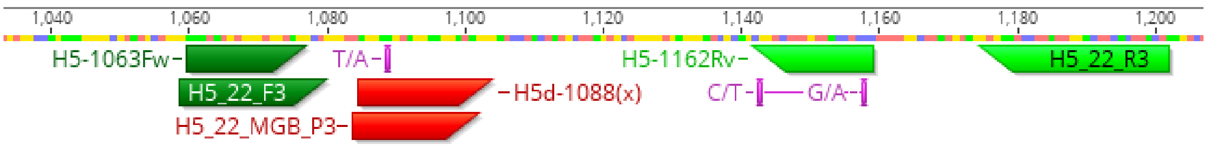


**Figure S1.** Primers and Probes used in H5 qPCR assay 1088/P3

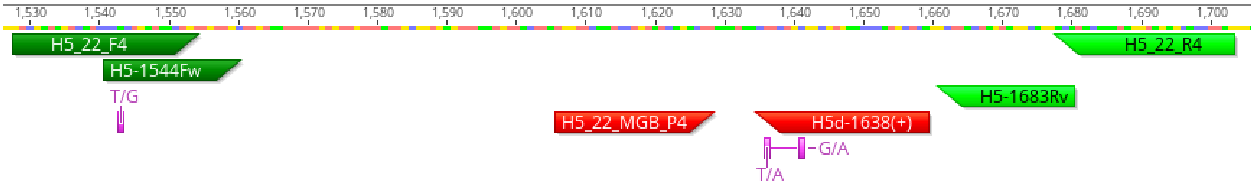


**Figure S2.** Primers and Probes used in H5 qPCR assay 1636/P4

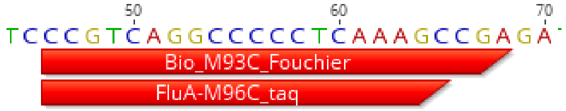


**Figure S3.** Published Influenza A (M segment, Fouchier) probe vs validated probe alignment

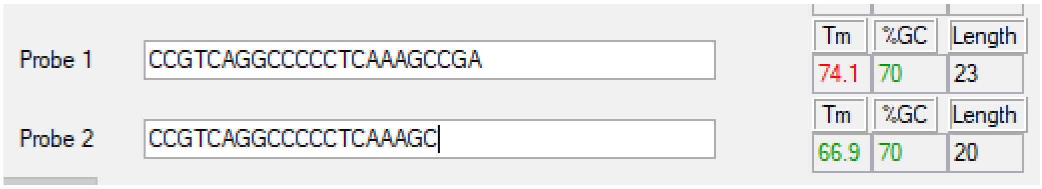


**Figure S4.** Published Influenza A (M segment, Fouchier) probe vs validated probe Primer Express Tm estimates.
